# Analysis of B-cell receptor repertoire to evaluate immunogenicity of monovalent Omicron XBB.1.5 mRNA vaccines

**DOI:** 10.1101/2024.01.22.24301315

**Authors:** Yohei Funakoshi, Kimikazu Yakushijin, Goh Ohji, Takaji Matsutani, Kazuhiko Doi, Hironori Sakai, Tomoki Sasaki, Takahiro Kusakabe, Sakuya Matsumoto, Yasuyuki Saito, Shinichiro Kawamoto, Katsuya Yamamoto, Taiji Koyama, Yoshiaki Nagatani, Keiji Kurata, Shiro Kimbara, Yoshinori Imamura, Naomi Kiyota, Mitsuhiro Ito, Hironobu Minami

**Author notes:** Corresponding author: Yohei Funakoshi, M.D., Ph.D. Division of Medical Oncology/Hematology, Department of Medicine, Kobe University Hospital and Graduate School of Medicine. Y.F., K.Yakushijin and G.O. contributed equally to this work.

## Abstract

Monovalent Omicron XBB.1.5 mRNA vaccines (BNT162b2 XBB.1.5 and mRNA- 1273.815) were newly developed and approved by the FDA in Autumn 2023 for preventing COVID-19. However, clinical efficacy for these vaccines is currently lacking. We previously established the Quantification of Antigen-specific Antibody Sequence (QASAS) method to assess the response to SARS-CoV-2 vaccination at the mRNA level using B-cell receptor (BCR) repertoire assay and the Coronavirus Antibody Database (CoV-AbDab). Here, we used this method to evaluate the immunogenicity of monovalent XBB.1.5 vaccines in healthy volunteers. We analyzed repeated blood samples before and after vaccination for the BCR repertoire to assess BCR/antibody sequences that matched SARS-CoV-2-specific sequences in the database. The number of matched unique sequences and their total reads quickly increased 1 week after vaccination. Matched sequences included those bound to the Omicron strain and Omicron XBB sublineage. The antibody sequences that can bind to the Omicron strain and XBB sublineage revealed that the monovalent XBB.1.5 vaccines showed a stronger response than previous vaccines or SARS-CoV-2 infection before the emergence of XBB sublineage. The QASAS method was able to demonstrate the immunogenic effect of monovalent XBB.1.5 vaccines for the 2023-2024 COVID-19 vaccination campaign.

## INTRODUCTION

In August 2021, the U.S. Food and Drug Administration (FDA) approved the first mRNA vaccines for severe acute respiratory syndrome coronavirus 2 (SARS-CoV-2), Pfizer-BioNTech BNT162b2 and Moderna mRNA-1273. These are monovalent mRNA vaccines against the original strain of SARS-CoV-2. These mRNA vaccines were highly effective in preventing symptomatic Coronavirus Disease 2019 (COVID-19) in the early phase of the pandemic.^1–3^ However, SARS-CoV-2 has undergone continuous evolution, leading to the emergence of novel variants of concern and high rates of breakthrough infection. In late 2022, because Omicron variants quickly dominated the global viral landscape, vaccine manufacturers accordingly developed original and Omicron BA.1 or BA.4/5 bivalent vaccines.^4–6^ In 2023, Omicron XBB sublineages became responsible for a large proportion of COVID-19. Because Omicron XBB sublineages efficiently evade immunity from previous infection or vaccines, monovalent Omicron XBB.1.5 mRNA vaccines (BNT162b2 XBB.1.5 and mRNA-1273.815) were newly developed and approved by the FDA in the Autumn of 2023.^7,8^

A few reports from *in vivo* studies suggest that monovalent XBB.1.5 vaccines provide promising activity in neutralizing emerging variants.^9^ Further, interim analyses of ongoing clinical trials have demonstrated the monovalent XBB.1.5 vaccines elicit potent neutralizing responses against not only variants of the omicron XBB sublineage (XBB.1.5, XBB.1.16, XBB.2.3.2, and EG.5.1) but also the recently emerged BA.2.86 variant.^8^ However, the clinical benefits of these effects are currently unknown, and further studies are required. Importantly, these XBB.1.5 vaccines are monovalent vaccines that do not contain vaccines against the original strain or other variants. Recently, concern has been expressed about the phenomenon of immune imprinting, in which exposure to a variant antigen following prior exposure to the primary antigen results in enhanced antibody production to the primary antigen and suppressed antibody production to the variant antigen.^10^ Given imprinting immunity by previous vaccination, monovalent XBB.1.5 vaccines might activate immunogenicity against the original strain and compromise the elicitation of antibodies against new SARS-CoV-2 variants.^7,8,10^

Previously, we established a new method to assess the response to SARS-CoV-2 vaccination at the mRNA level by quantifying antigen-specific antibody sequences, named the “Quantification of Antigen-specific Antibody Sequence (QASAS) method”.^11^ This method uses B-cell receptor (BCR) repertoire data on activation of humoral immunity and a database of BCR sequences that bind to target antigens, such as the Coronavirus Antibody Database (CoV-AbDab).^12^ Since the CoV-AbDab contains information on variants to which each antibody sequence binds, our method allows us to infer the antigen specificity of antibody sequences generated by the vaccine. In this study, we used the QASAS method to evaluate immunogenicity after monovalent Omicron XBB.1.5 m RNA vaccination.

## MATERIAL AND METHODS

### Participants

Immune response following monovalent Omicron XBB.1.5 mRNA vaccination (BNT162b2 XBB.1.5 Pfizer-BioNTech] and mRNA-1273.815 [Moderna]) was evaluated in three healthy volunteers enrolled in October 2023. Two volunteers received BNT162b2 XBB.1.5 as the sixth vaccination, and one volunteer received mRNA-1273.815 as the seventh vaccination. All participants had no previous history of COVID-19 and were confirmed to be negative for antibodies against SARS-CoV-2 nucleocapsid protein using a QuaResearch COVID-19 Human IgM IgG ELISA Kit (nucleocapsid protein) (Cellspect, Inc., RCOEL961-N, Iwate, Japan). Blood samples were chronologically collected pre- and post-vaccination.

The study protocol was approved by Kobe University Hospital Ethics Committee (No. B2356701). The study was conducted in accordance with the principles of the Declaration of Helsinki. The participants provided written informed consent for this research.

### Sample collection and processing

Peripheral blood samples were collected using heparin-containing tubes. Peripheral blood mononuclear cells (PBMCs) were isolated using Ficoll-Paque Plus (GE Healthcare, Little Chalfont, UK) and stored with CELLBANKER (ZENOGEN PHARMA, Fukushima, Japan) at -80°C until analysis. Total RNA was extracted with TRIzol LS (Thermo Fisher Scientific, Waltham, MA, USA) from PBMC for B cell receptor (BCR) repertoire analysis and purified with an RNeasy Mini Kit (Qiagen, Hilden, Germany) in accordance with the manufacturer’s instructions. RNA amounts and purity were measured with an Agilent 2200 TapeStation (Agilent Technologies, Santa Clara, CA, USA).

Serum samples were obtained by centrifuging blood samples for 10 min at 1000 × g at room temperature, and immediately transferred to a freezer kept at −80 °C.

### Measurement of SARS-CoV-2 strain-specific IgG antibody titers

The SARS-CoV-2 strain-specific anti-spike (anti-S) IgG titers were measured with in-house ELISA using trimeric full-length spike proteins generated by a silkworm-baculovirus expression system, as previously described.^13^ Briefly, purified target proteins of the WT and Omicron (BA.1) strains were used as coating antigens for ELISA. Optical density at 450 nm (OD 450 nm) and 570 nm (OD 570 nm) was read using a microplate reader after ELISA, and the anti-S IgG titers of each sample were reported as the OD value (OD 450 nm − 570 nm).

With regard to antibody titers against nucleocapsid proteins, these were measured with a QuaResearch COVID-19 Human IgM IgG ELISA Kit (nucleocapsid protein) (Cellspect, Inc., RCOEL961-N). This kit detects antibody titers based on the indirect ELISA method and comes with different immobilized antigenic proteins. The plate of the ELISA kit (nucleocapsid protein) was immobilized with a recombinant nucleocapsid protein (full length) of SARS-CoV-2 expressed in *Escherichia coli*. Nucleocapsid protein in serum samples was measured in accordance with the manufacturer’s measurement protocol. Optimal OD cut-off values of anti-S1 IgG antibody and anti-nucleocapsid IgG antibody for seroconversion were determined to be 0.7.^14^

### B-cell receptor repertoire analysis

BCR repertoire analysis was performed using unbiased next-generation sequencing developed by Repertoire Genesis, Inc. (Osaka, Japan).^15^ Briefly, cDNA was synthesized from total RNA using the polyT18 primer (BSL-18E) and Superscript III reverse transcriptase (Invitrogen, Carlsbad, CA, USA). After synthesizing double-strand (ds)- cDNA, the P10EA/P20EA dsDNA adaptor was ligated and cut with the *Not*I restriction enzyme. Nested PCR was performed with KAPA HiFi DNA Polymerase (Kapa Biosystems, Woburn, MA, USA) using IgG constant region-specific primers (CG1 and CG2) and P20EA. The amplicon library was prepared by amplification of the second PCR products using P22EA-ST1 and CG-ST1-R. Index (barcode) sequences were added by amplification with a Nextera XT Index Kit v2 Set A (Illumina, San Diego, CA, USA). Sequencing was performed using the Illumina MiSeq paired-end platform (2×300 bp). BCR sequences were assigned based on identity with reference sequences from the international ImMunoGeneTics information system® (IMGT) database (http://www.imgt.org) using repertoire analysis software originally developed by Repertoire Genesis, Inc. (Osaka, Japan).

### Database and SARS-CoV-2-specific sequence search

BCR/antibody sequences specific for SARS-CoV-2 were downloaded from CoV-AbDab (http://opig.stats.ox.ac.uk/webapps/covabdab/). Data updated on 13 June 2023, containing 12,536 entries, were used as reference. Antibodies with immunoglobulin heavy chain sequences with identical V and J genes and CDR3 amino acid sequences in the CoV-AbDab database were grouped as unique reference sequences. These unique reference sequences were classified based on their binding specificity to viruses, strains, and sublineages. Sequence classification was performed by matching virus search string (SARS-CoV-2), strain search strings (WT, Alpha, Beta, Gamma, Delta, Epsilon, Iota, Kappa, Lambda, Mu, Omega, Omicron) or sublineage search strings (BA1, BA2, BA3, BA4/5, BA5, BQ1, BR2, CA1, BM111, XBB) against the database registration information. Sequences detected in BCR repertoire analysis of PBMCs from vaccinated patients were searched against the classified reference sequence, and sequences with CDR3 amino acid sequence exact match (Levenshtein distance 0, LV0) or 1 amino acid mismatch (LV1) were counted.^11^

## RESULTS

### Serological outcomes after monovalent Omicron XBB.1.5 mRNA vaccination on anti-SARS-CoV-2 strain-specific spike IgG antibody measurement by ELISA

The timelines of vaccination for participants are described in Figure 1A. Anti-spike IgG antibody titers for original and Omicron strain were measured pre- and 14 days after monovalent XBB.1.5 vaccination in all participants, and titers were increased after the vaccination (Figure 1B).

**Figure 1.**
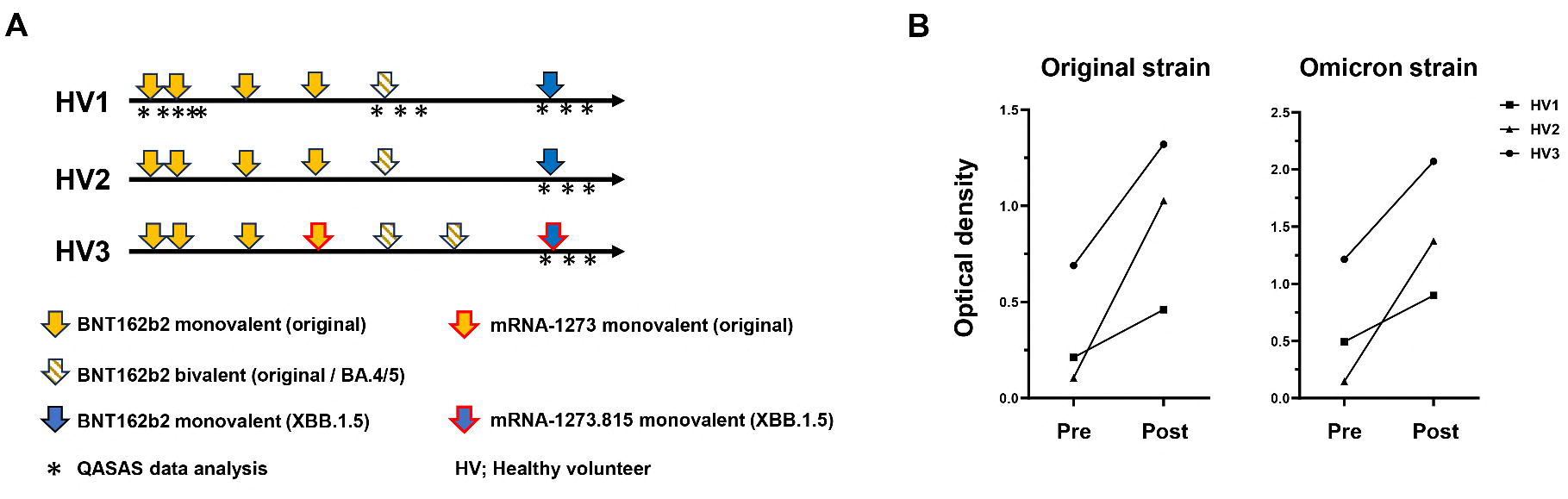
Schedule of vaccination and blood sampling for QASAS analysis and measurement of each anti-spike antibody. (A) Arrows indicate vaccinations, and asterisks indicate blood sample collection for QASAS analysis. Three healthy volunteers (HV1, HV2 and HV3) received the monovalent Omicron XBB.1.5 mRNA vaccines (BNT162b2 XBB.1.5 or mRNA-1273.815) in Autumn, 2023. (B) Anti-spike antibody of original and Omicron strain titers was measured before and after administration of the monovalent Omicron XBB.1.5 mRNA vaccines by fully automated commercial immunoassays in the three participants.

### BCR repertoire data for the QASAS method

To evaluate the immunogenicity of vaccines, we analyzed BCR repertoire data pre- and post-vaccination using BCR sequences that bind to SARS-CoV-2 (QASAS method) (Figure 2A).

**Figure 2.**
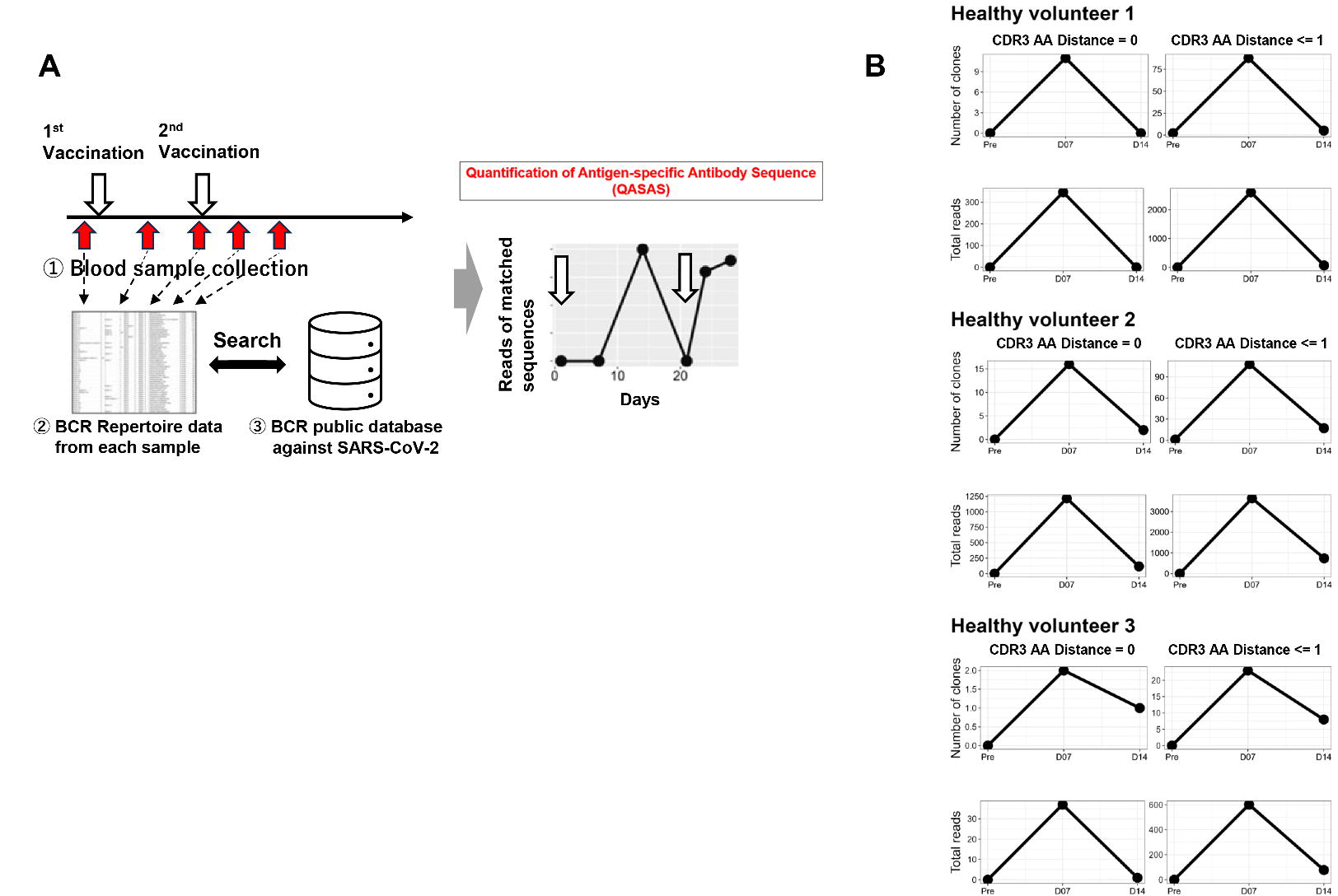
Immunogenicity of monovalent Omicron XBB.1.5 mRNA vaccines evaluated by the Quantification of Antigen-specific Antibody Sequence (QASAS) method. (A) Schema of the QASAS method. We collected blood samples over time pre- and post- vaccination to analyze the BCR repertoire. We then analyze the extent to which BCR sequences in the database were contained in the obtained BCR sequence data set, and analyzed the transition of matched BCR sequences. (B) Changes over time in the number (Unique Read, upper) and total reads (Total Read, lower) of SARS-CoV-2- specific sequences following vaccination. Before and after vaccination (Day 0, Day 7 and Day 14 after vaccination), SARS-CoV-2-specific sequences were retrieved from the BCR repertoire data. LV, Levenshtein.

Nine blood samples from three participants (3 samples for each participant: pre-, and 7 and 14 days after vaccination) were analyzed for BCR repertoire. A total of 1,097,175 sequences were obtained, of which 1,072,068 in-frame sequences with V and J genes and CDR3 amino acid assignments were used. We then used the QASAS method to analyze the extent to which SARS-CoV-2-specific sequences were included in the BCR sequence data set obtained from each blood sample of XBB-vaccinated donors (Figure 2B). Although no matched sequences were detected before vaccination, the number of matched unique sequences and total reads quickly increased 1 week after vaccination and decreased thereafter (Figure 2B). The QASAS method clearly demonstrated the response to monovalent XBB.1.5 vaccination.

### Strains and sublineages to which each matched BCR/antibody sequence bound

We analyzed which strains and sublineages were bound by each matched sequence, and calculated the number of unique matched sequences along with their total reads for each strain and sublineage (Figure 3). Although many of the matched sequences were antibody sequences that bound to wild-type (original strain), some of which bound to the Omicron strain and XBB sublineage among Omicron strain were also detected (Figure 3). All matched sequences peaked in the first week after vaccination regardless of strain and sublineage.

**Figure 3.**
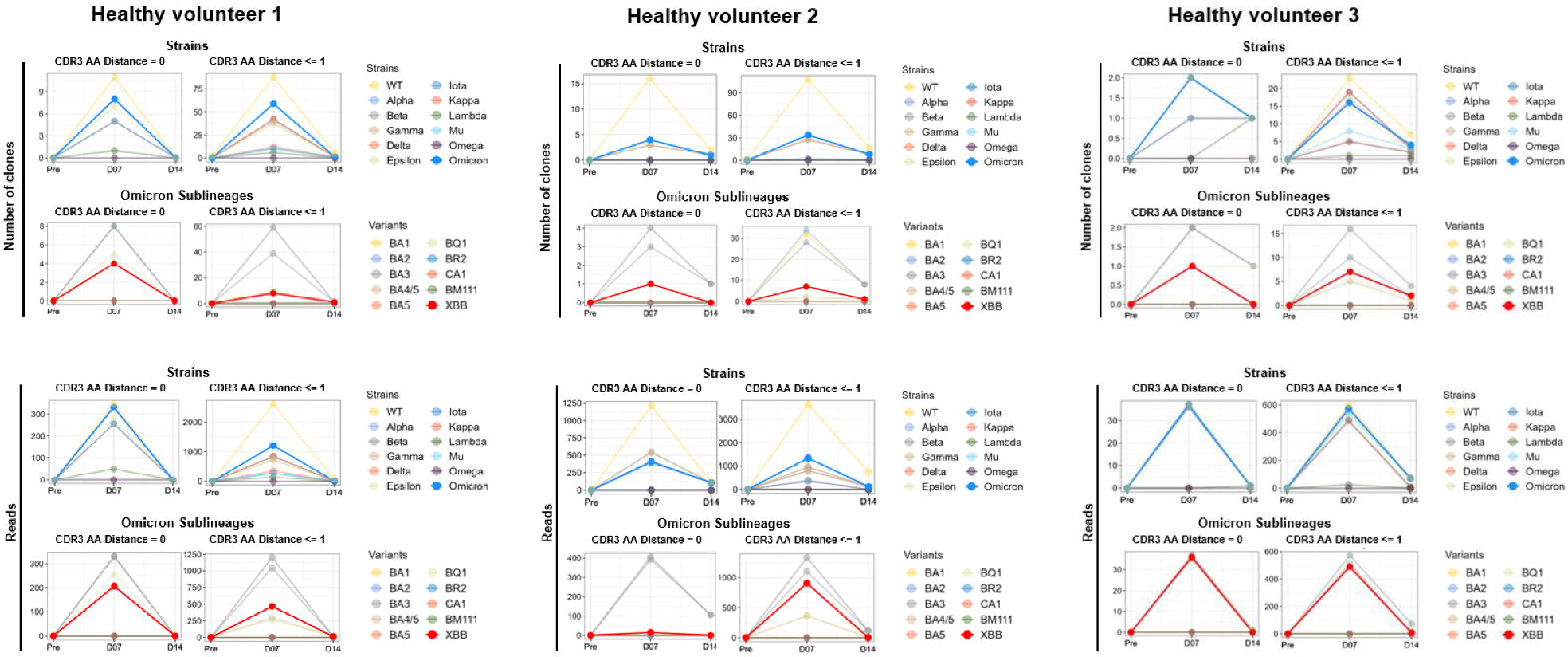
Analysis of strains and sublineages to which the matched BCR/antibody sequences bound. Strains and sublineages and the matched sequences which bound to them were analyzed, and the number of matched unique sequences and reads for each strain and sublineage were counted. Binding specificities for 12 SARS-CoV-2 strains (WT, Alpha, Beta, Gamma, Delta, Epsilon, Iota, Kappa, Lambda, Mu, Omega, and Omicron) and 10 Omicron sublineages (BA1, BA2, BA3, BA4/5, BA5, BQ1, BR2, CA1, BM111, and XBB) in the database were searched by search string.

### Comparison of immune responses with monovalent Omicron XBB.1.5 mRNA vaccines and other antigens (previous vaccines or SARS-CoV-2 infection before XBB.1.5 emergence)

In healthy volunteer 1, evaluation of vaccine-induced immune responses by QASAS has so far been carried out not only for the sixth vaccine (monovalent XBB.1.5 vaccine) but also for the first, second, and fifth vaccines (Figure 1A). Compared with previous vaccines (monovalent original mRNA vaccines and bivalent original/omicron BA.4/5 mRNA vaccine), monovalent XBB.1.5 vaccine tended to induce more total reads of matched sequences that bind to the Omicron strain and XBB sublineage (Figure 4A).

**Figure 4.**
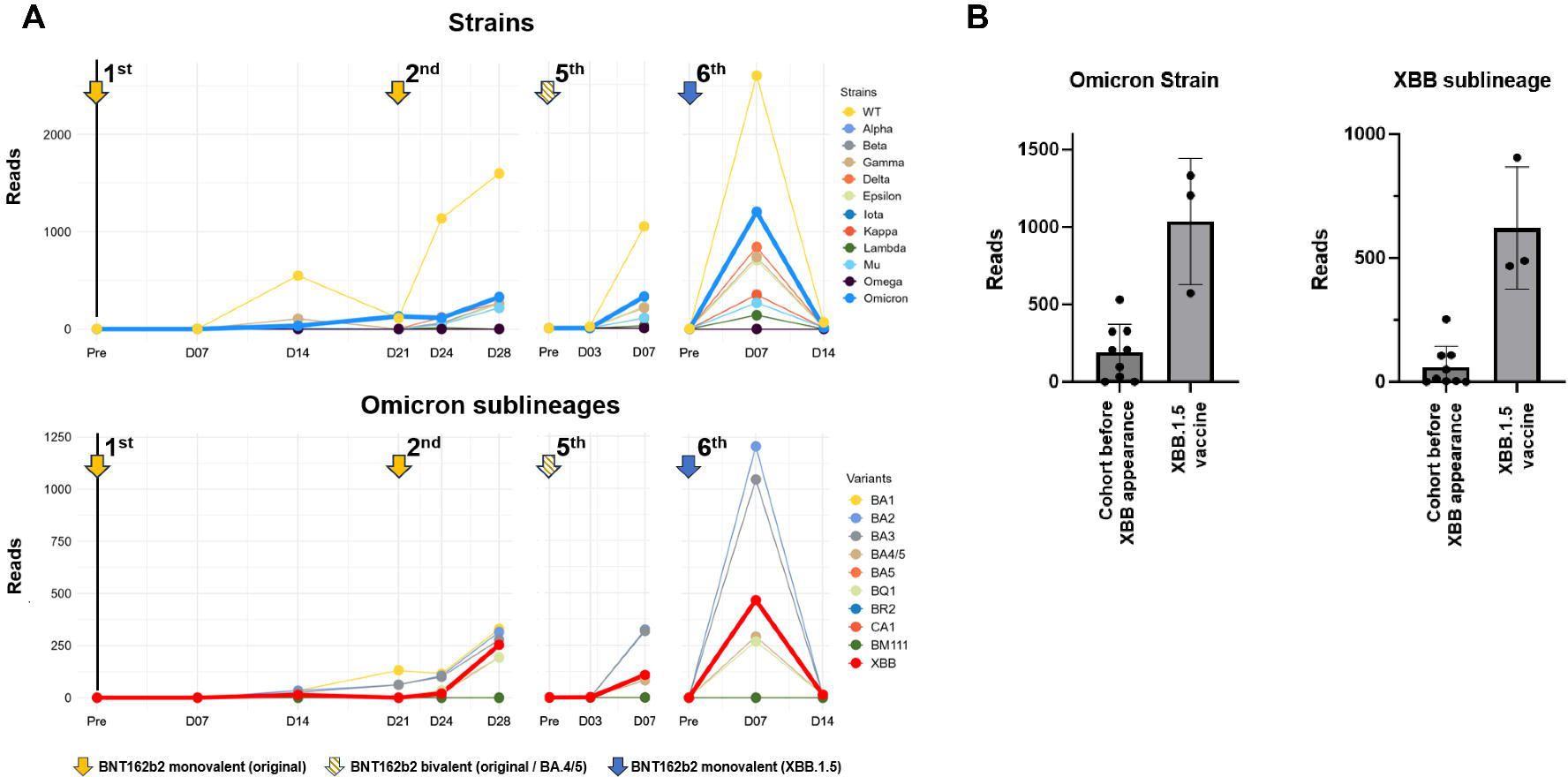
Comparison of immune responses with monovalent Omicron XBB.1.5 mRNA vaccines and wild-type antigens (vaccines or infection) (A) Changes over time in reads of SARS-CoV-2 strains and Omicron sublineage-specific sequences (CDR3 AA Distance ≤ 1) after each vaccination in healthy volunteer 1 (HV1). The schedule for vaccinations and QASAS analysis is shown in Figure 1A, HV1. (B) Three cases of SARS-CoV-2 infection in early 2020 and six cases of individuals who had received previous vaccines (monovalent original mRNA vaccines and bivalent original/Omicron BA.4/5 mRNA vaccine) were analyzed using the QASAS method. In reads of Omicron XBB sublineage-specific sequences (CDR3 AA Distance ≤ 1), these nine cases before the appearance of Omicron XBB sublineage were compared with current monovalent XBB.1.5 vaccine recipients.

In our previous research, we evaluated the immune response in individuals primed with previous vaccines or SARS-CoV-2 infection through the QASAS method.^11^ These included three cases of SARS-CoV-2 infection in early 2020 and six cases of individuals who had received previous vaccines (monovalent original mRNA vaccines and bivalent original/Omicron BA.4/5 mRNA vaccine). These nine cases were treated as a cohort exposed to antigens before the appearance of the Omicron XBB sublineage (Supplementary Table 1). We then compared their QASAS data with that of the current monovalent XBB.1.5 vaccine recipients. The total reads of matched sequences binding to the XBB sublineage tended to be higher after monovalent XBB.1.5 vaccination (Figure 4B).

## DISCUSSION

We found that the QASAS method detected immune response to monovalent XBB.1.5 vaccines. In this study, we evaluated immunogenicity in three healthy volunteers after monovalent Omicron XBB.1.5 mRNA vaccination. Regardless of strain or sublineage, B-cell response quickly increased 1 week after vaccination and then quickly decreased (Figures 2B and 3). Importantly, focusing on antibody sequences that can bind to the Omicron strain and XBB sublineage, the monovalent XBB.1.5 vaccines showed a stronger response than the previous vaccines and SARS-CoV-2 infection before the emergence of the XBB sublineage.

The QASAS method has several advantages over the measurement of antibody titers by conventional serological tests in determining vaccine response. Because CoV-AbDab contains binding information of BCR/antibody sequences to specific strains and sublineages of SARS-CoV-2, the QASAS method provides this information for each individual matched BCR/antibody sequence. Since information on new strains and sublineages are being continuously added to this database, it is possible to examine whether BCR/antibody sequences induced by previous vaccinations can work for newly emergent variants if the BCR repertoire data are preserved. Furthermore, compared to conventional tests, which detect antibodies with a long half-life, the QASAS method directly detects BCR mRNA as short-lived as B cell activity, making it possible for our method to monitor the real-time activity of humoral immunity. Given that the initial vaccine immunization in our previous study took two weeks to increase the SARS-CoV- 2-specific BCR/antibody sequences,^11^ we consider that the rapid peak in B-cell activation after antigen exposure indicates the effect of a boosted immune response.

A few clinical trials of the efficacy of monovalent XBB.1.5 vaccines have been reported.^8,16^ Results showed that monovalent XBB.1.5 vaccines elicited potent neutralizing responses against variants of the omicron XBB-lineage (XBB.1.5, XBB.1.6, XBB.2.3.2) and more recent (EG.5.1, FL.1.5.1) variants.^8,16^ Further, these vaccines also increased neutralizing antibody responses against various strains and sublineages, including the original strain.^8,16^ This outcome has been attributed to immune imprinting – namely, exposure to a variant antigen following exposure to a primary antigen results in increased antibody production to the primary antigen than to the variant antigen.^16^ Accordingly, given the impact of immune imprinting, our detection of antibody sequences against various strains and sublineages following monovalent XBB.1.5 vaccination appears reasonable.

Immunological imprinting is considered to suppress antibody production against the XBB sublineage even after monovalent XBB.1.5 vaccination,^17^ and a research group in fact demonstrated the impact of imprinted immunity in an experimental animal model.^10^ However, another research group demonstrated that monovalent XBB.1.5 booster vaccination promoted higher neutralizing antibody titers against the XBB sublineage than a booster vaccination by previous vaccines (original monovalent or original/BA.4/5 bivalent) in an animal model.^9^ In our study, monovalent XBB.1.5 vaccination elicited more antibody binding to the XBB sublineage than prior vaccination or SARS-CoV-2 infection before XBB.1.5 emergence, supporting the validity of monovalent XBB.1.5 vaccination.

The major limitation of this study is that only three healthy volunteers were evaluated. Confirmation of our early findings therefore requires further research using a larger sample size. Further, although we compared the immune response induced by exposure to previous antigens other than the XBB sublineage with that of exposure to monovalent XBB.1.5 vaccine, a comprehensive understanding of immune imprinting requires the comparison of cases receiving the monovalent XBB.1.5 vaccine as a booster vaccination with those receiving the vaccine as first-time immunization. This would be difficult, however, as the number of individuals with no history of either vaccination or SARS-CoV-2 infection is limited. Finally, matched BCR/antibody sequences in CoV-AbDab might not necessarily be useful for protecting against infection in clinical practice. However, the QASAS method revealed that there was an increase in specific sequences in the first week after vaccination and a subsequent decrease, which suggests that these antibody sequences are indeed induced by vaccination and play a role in immune defense against various strains and sublineages.

In conclusion, we were able to evaluate the immunogenicity of monovalent XBB.1.5 mRNA SARS-CoV-2 vaccines by our unique QASAS method, which uses a BCR repertoire and database. The XBB vaccines are expected to induce the rapid production of antibodies capable of binding to Omicron XBB.

## Author contributions

Designed research, Y.F., K.Yakushijin, G.O., T.M. and H.M.; performed research, Y.F., K.Yakushijin, G.O., K.D., H.S., T.S., S.M., Y.S., K.Yamamoto, T.Koyama., Y.N., K.K., S.Kmbara., Y.I., N.K. and H.M; contributed vital new analytical tools, T.M., K.D., H.S., T.S. and T.Kusakabe; analyzed data, Y.F. K.Yakushijin, G.O., T.M., K.D., H.S., T.S. and T.Kusakabe; and wrote the paper, Y.F. K.Yakushijin, G.O., T.M. S,M., Y.S., S.Kawamoto, K.Yamamoto, T.koyama, Y.N., K.K., S.Kimbara, Y.I., N.K., M.I. and H.M.

## Ethics statement

This study was conducted in accordance with the principles of the Declaration of Helsinki.

## Participat consent statement

All participants provided written informed consent for this research.

## Clicical trial registraion number

The study protocol was approved by Kobe University Hospital Ethics Committee (No. B2356701).

## Correspondence

Yohei Funakoshi, Division of Medical Oncology/Hematology, Department of Medicine, Kobe University Hospital and Graduate School of Medicine, Kobe, Japan

## Data Availability

All data produced in the present study are available upon reasonable request to the authors.

