## Supplemental Table 1 for "Analysis of B-cell receptor repertoire to evaluate immunogenicity of monovalent Omicron XBB.1.5 mRNA vaccines"

Supplementary Table 1

|  | **Types of antigen exposure** | **Date of**  **QASAS analysis** | **Note** |
| --- | --- | --- | --- |
| No.1 | Infection | Sep/2020 | First infection in non-vaccinated individual |
| No.2 | Infection | Sep/2020 | First infection in non-vaccinated individual |
| No.3 | Infection | Sep/2020 | First infection in non-vaccinated individual |
| No.4 | 1^st^ Vaccination (monovalent BNT162b2) | Apr/2021 | Healthy volunteer |
| No.5 | 2^nd^ Vaccination (monovalent BNT162b2) | May/2021 | Healthy volunteer |
| No.6 | 5^th^ Vaccination (bivalent BNT162b2) | Nov/2023 | Healthy volunteer |
| No.7 | 4^th^ Vaccination (monovalent mRNA-1273) | Sep/2022 | Healthy volunteer |
| No.8 | 4^th^ Vaccination (bivalent BNT162b2) | Jun/2023 | Patient with haematological malignancy |
| No.9 | 1^st^ Vaccination (monovalent BNT162b2) | Dec/2022 | Patient with haematological malignancy |

Cohort exposed to antigens before appearance of the Omicron XBB sublineage
